# Assessing Adequacy and Variability in Semi-Structured Ga-68 PSMA PET/CT Reports for Prostate Cancer in view of auto-input to AI models

**DOI:** 10.1101/2025.04.10.25325625

**Authors:** Jagrati Chaudhary, Anil Kumar Pandey, Param Dev Sharma, Sanjay Kumar, Kunhiparambath P Haresh, Rakesh Kumar

**Author notes:** **Corresponding Author:** Dr Rakesh Kumar Address of Correspondence: **Anil Kumar Pandey** Professor Department of Nuclear medicine, All India Institute of Medical Sciences, Ansari Nagar, New Delhi-110029, India. mail.

## Abstract

**Introduction:** Gallium-68 Prostate specific membrane antigen (Ga-68 PSMA) PET/CT has significantly improved prostate cancer imaging by offering superior sensitivity and specificity over conventional modalities. However, the effectiveness of this diagnostic tool depends on the quality and consistency of reporting. This study evaluates the adequacy, consistency, and AI compatibility of semi-structured Ga-68 PSMA PET/CT reports.

**Methods:** Essential reporting elements were determined through consensus among nuclear medicine physicians, urologists, and radiotherapists. Two hundred Ga-68 PSMA PET/CT reports from prostate cancer patients (January 2020–June 2024) were analysed. Reporting Adequacy Score (RAS) assessed the percentage of clinical needs met, while Variability Index (VI) quantified inconsistencies in terminology. Statistical analyses, including descriptive statistics, frequency distribution, and boxplots, were performed to evaluate reporting trends.

**Results:** RAS ranged from 31% to 78%, with 91% of reports being partially adequate (50– 80% RAS) and 9% inadequate (<50% RAS). Key clinical details, such as Lesion Maximum Standardized Uptake Value (SUVmax), Neurovascular bundle involvement, and Bone metastasis lesion count, were frequently missing. Inconsistent terminology was observed in lesion descriptions, lymph node involvement, and uptake patterns, with VI ranging from minimal (4%) to high (67%). Reports with lower RAS and high VI were less suitable for AI-based data extraction, posing challenges to automation.

**Conclusion:** The majority of Ga-68 PSMA PET/CT reports were partially adequate, with significant missing details and considerable variability in terminology. The variability was evenly distributed across minimal, moderate, and high levels. Training an AI model on such reports would likely result in slower learning and compromised performance.

## Introduction

Prostate cancer is a major global health concern, ranking as the second most common malignancy in men and a leading cause of cancer-related mortality. With approximately 1.4 million new cases diagnosed annually and over 375,000 deaths worldwide, effective diagnostic and management strategies are essential for improving patient outcomes [1]. Accurate staging and early detection of disease recurrence or progression play a crucial role in the successful management of prostate cancer [2].

Ga-68 PSMA PET/CT has transformed prostate cancer diagnosis by providing unparalleled sensitivity and specificity compared to traditional imaging techniques such as CT, MRI, and bone scintigraphy [3]. This advanced imaging modality excels in detecting primary prostate tumors, nodal involvement, and distant metastases. Moreover, when integrated with artificial intelligence (AI), it offers even greater diagnostic precision [4].

In spite of the above advancements, the clinical utility of Ga-68 PSMA PET/CT is significantly influenced by how findings are conveyed in nuclear medicine reports [5]. At our center, a high-volume tertiary healthcare institution, Ga-68 PSMA PET/CT scans for prostate cancer patients are frequently requested by urologists, oncologists, and radiotherapists. These reports typically follow a structured format with sections such as Indications, Head and Neck, Thorax, Abdomen-Pelvis, and Musculoskeletal System. However, within each section, findings are described in free-text format, which can lead to inconsistencies in terminology. Such variability poses challenges for AI systems that rely on natural language processing (NLP) to extract meaningful information from these reports [6].

Several key questions remain unanswered regarding the effectiveness of Ga-68 PSMA PET/CT reports issued for prostate cancer patients: To what extent do these reports address the clinical needs of stakeholders, how much variability exists in the terminology used, and Is the report structure optimized for seamless AI processing (in this context the word “seamless” means the structure and content of the reports are fully optimized for efficient, error-free, and automated AI interpretation without requiring additional pre-processing, corrections, or manual intervention. )? This study aims to answer these questions by analysing Ga-68 PSMA PET/CT reports issued for prostate cancer patients.

## Materials and Methods

### Study Design and Data Collection

This retrospective study was conducted at a tertiary care hospital. Ethical approval for the study was obtained from the institutional ethics committee, ensuring adherence to established ethical research guidelines [A2193/26.09.2024]. The study included 200 patients who underwent Ga-68 PSMA PET/CT scans between January 2020 and June 2024.

The clinical reports of Ga-68 PSMA PET/CT study of patients according to the following inclusion and exclusion criteria were analyzed.

### Inclusion Criteria

- Patients with histologically confirmed prostate cancer.
- Patients who underwent Ga-68 PSMA PET/CT imaging for baseline evaluation.
- Clinical reports issued by the facility is available in the departmental records.
- Reports generated within a time period between January 2020 and June 2024.

### Exclusion Criteria

- Patients with missing histopathology report.
- Patients who did not provide informed consent for the use of their medical records in research.
- Reports written entirely in a free-text format without structured components.

### Defining Essential Information in Ga-68 PSMA PET/CT Reports

To address the question: *“To what extent do Ga-68 PSMA PET/CT reports issued to prostate cancer patients address the clinical needs of stakeholders?”* it was essential to first establish a standardized list of information to be assessed in each report. The following procedure was adopted to create this list:

1. **Literature Review** – A comprehensive review of existing literature was conducted to identify the essential information that should be included in a Ga-68 PSMA PET/CT report [7, 8]. Based on this review, a preliminary draft was created.
2. **Consultation with a Radiotherapist** – The preliminary draft was reviewed and discussed with a radiotherapy consultant. The clinical needs from a radiotherapist’s perspective were incorporated, leading to the development of the second draft.
3. **Consultation with a Urologist** – The second draft was then discussed with a urologist, and their input on clinical requirements from a urology perspective was integrated into the third draft.
4. **Review by a Nuclear Medicine Physician** – A nuclear medicine (NM) physician reviewed the third draft and provided further refinements, resulting in the fourth draft.
5. **Consensus Among Stakeholders** – The fourth draft was circulated among all key stakeholders, including NM physicians, urologists, and radiotherapists. Their collective feedback was incorporated, and a consensus was reached to finalize the agreed-upon list of essential attributes to be searched in each report (**Table 1**).

**Table 1:**
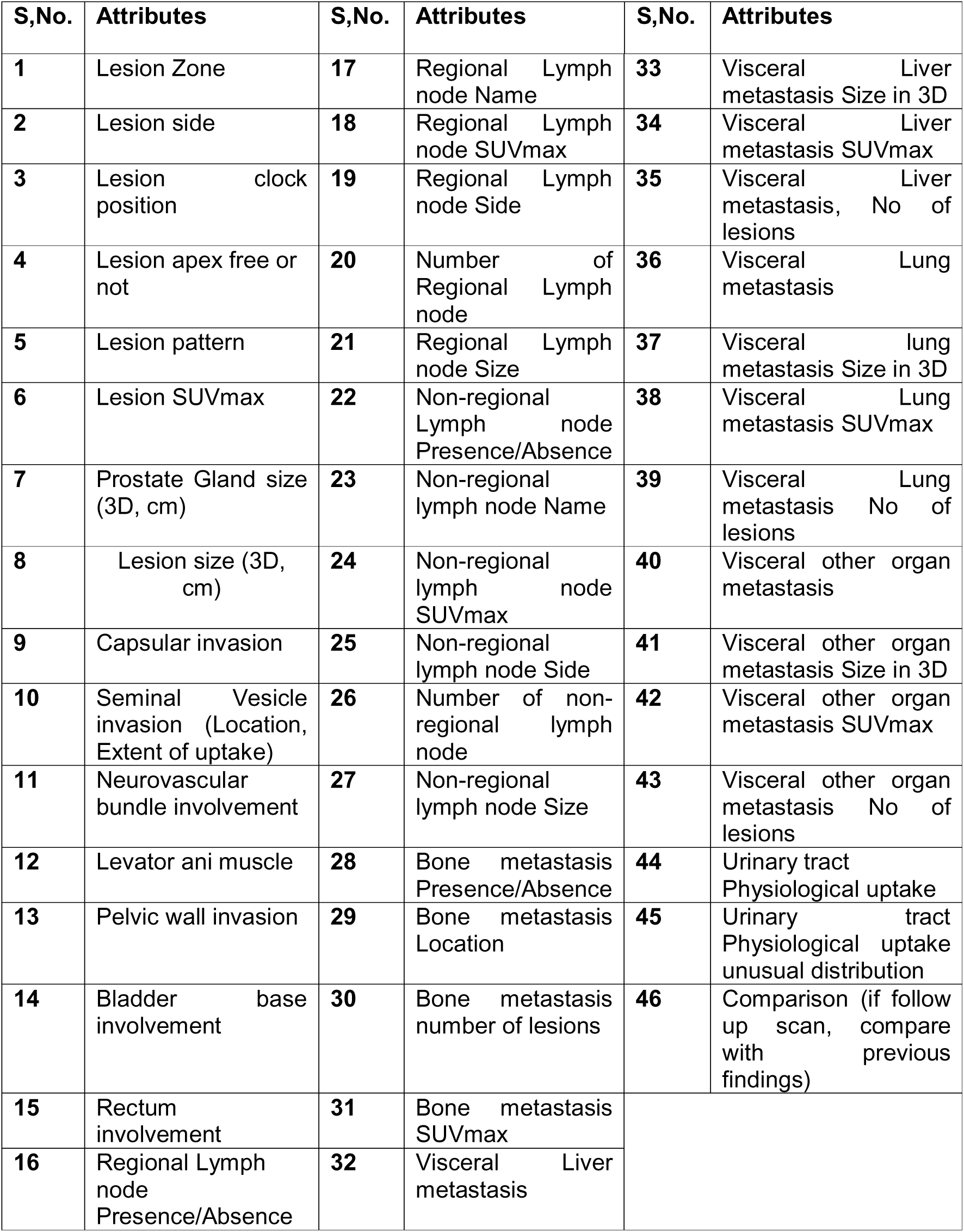
List of essential attributes to be searched in each report.

From now onwards, we will use **“Reporting Adequacy Score”** as a metric for **“Percentage of Clinical Needs Were Answered”.**

**Reporting Adequacy Score (RAS)** is a quantitative metric that evaluates the extent to which Ga-68 PSMA PET/CT reports address essential clinical needs. It is calculated as the percentage of predefined essential attributes present in each report, reflecting the adequacy of reporting in terms of completeness and clinical relevance.

### Formula for RAS Calculation

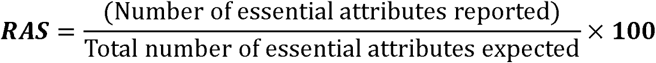

where:

- **Essential attributes reported** = The number of clinically relevant attributes documented in a given report.
- **Total essential attributes expected** = The total number of predefined attributes that should ideally be included in the report.

### Interpretation of RAS

- **100% RAS** → The report is fully adequate, covering all necessary clinical attributes.
- ≥**80% RAS** → The report is mostly complete but may lack some details.
- **50-80% RAS** → The report is partially adequate, with notable missing elements.
- **<50% RAS** → The report is inadequate and may not meet clinical needs effectively.

### Assessing Variability in Terminology Used for Essential Attributes

While manually reviewing the reports for the presence of essential attributes and their corresponding values, particular attention was given to how these attributes were described across different reports. Any variations in terminology used to convey the same attribute were carefully documented. The exact, wording found in each report was recorded until all 200 reports had been reviewed. These variations were then compiled into a frequency table, categorizing the different terminologies used to express the attributes and their values.

The **Variability Index (VI)** is a metric used to quantify the inconsistency in reporting practices. It measures how frequently different terminologies or descriptions are used for the same attribute across multiple reports. A higher variability index indicates greater inconsistency.

### Formula for Variability Index (VI)

The **Variability Index** for a given attribute can be calculated as:

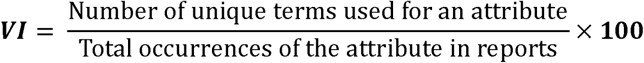

where:

- **Number of unique terms** = Different terminologies used to describe the same clinical attribute.
- **Total occurrences** = Total times the attribute was reported in the dataset.

### Interpretation of Variability Index

- **VI = 0%** → Fully standardized (only one term used).
- **VI < 10%** → Minimal variability (mostly consistent terminology).
- **VI 10-30%** → Moderate variability (some inconsistencies in wording).
- **VI > 30%** → High variability (significant inconsistency, affecting AI-readiness).

### Evaluating AI Optimization of Report Structure

AI models could use imaging data and its computed derivatives as well as clinical reports to enhance diagnosis, staging, and prognostication. Such models often prioritize or focus on three key areas: lesion detection and segmentation, automated reporting, and prognostic analytics.

Whereas computed derivatives from image-data may be the basis of Deep learning models that identify and segment lesions in Ga-68 PSMA PET/CT scans, report-text could also be utilized to extract clinical parameters. The later approach may improve the epistemic transparency by making outcomes better verifiable for the user, and associated NLP performance gets better if reports are well structured.

We are developing an AI model for prostate cancer diagnosis, local staging, and prognostication that would use data from Ga-68 PSMA PET/CT scans, histopathology reports, and other clinical modalities. Auto extraction of information from report-text is a planned input along side imaging data and its computed derivatives. Structured reports facilitate the auto-extraction of information with lower margins of error as compared to semi-structured or poorly structured reports. Although AI can work with available data, structured reporting would boost the performance by shortening the training loop and enhancing learning robustness. Further structured reports can facilitate end-user level comparison of AI-model outcomes with other traditional models.

Since several AI models give results but are hardly explainable, end-user acceptability will remain a challenge. Reports with a high RAS score can address the issue of making AI outcomes better acceptable, whereas high VI reports besides making AI-driven data extraction harder also lowers the end-user’s acceptability of outcomes. Standardized reporting improves AI readiness and supports better understanding of the reliability and accuracy of the model involved.

### Statistical Analysis

The age, Gleason score, and PSA levels of the patients were recorded. All 200 Ga-68 PSMA PET/CT reports were manually reviewed line by line to identify whether the attributes defined by stakeholders were present. If present, the various terminologies used for these attributes, along with the specific words, phrases, or sentences describing their values, were documented. These observations were entered into Microsoft Excel Spreadsheet for further analysis.

Reporting Adequacy Score (RAS) and Variability Index (VI) were calculated. The data in Excel were analysed and presented using descriptive statistics. MATLAB R2021a was used for generating frequency tables, boxplots, and other graphical representations.

## Results

Majority prostate cancer patients who underwent Ga-68 PSMA PET/CT scan were within the typical age range: 66.6 ± 7.9 (mean ± 1 S.D.) years. 126 out of 200 patients reports (63%) had documented Gleason scores, with a mean Gleason score of 7.54 ± 0.86 indicating that most of the patients had intermediate- to high-risk prostate cancer. Prostate-specific antigen (PSA) levels were documented in 84% (168 out of 200) of the reports available with a mean PSA level of 188.68 ng/dL and having wide range spanning from 0.02 to 13,506 ng/dL suggesting a highly variable datasets encompassing both early and advanced-stage prostate cancer cases. The box plots showing distribution of age, PSA levels and Gleason score is shown in **Figure-1**.

**Figure 1:**
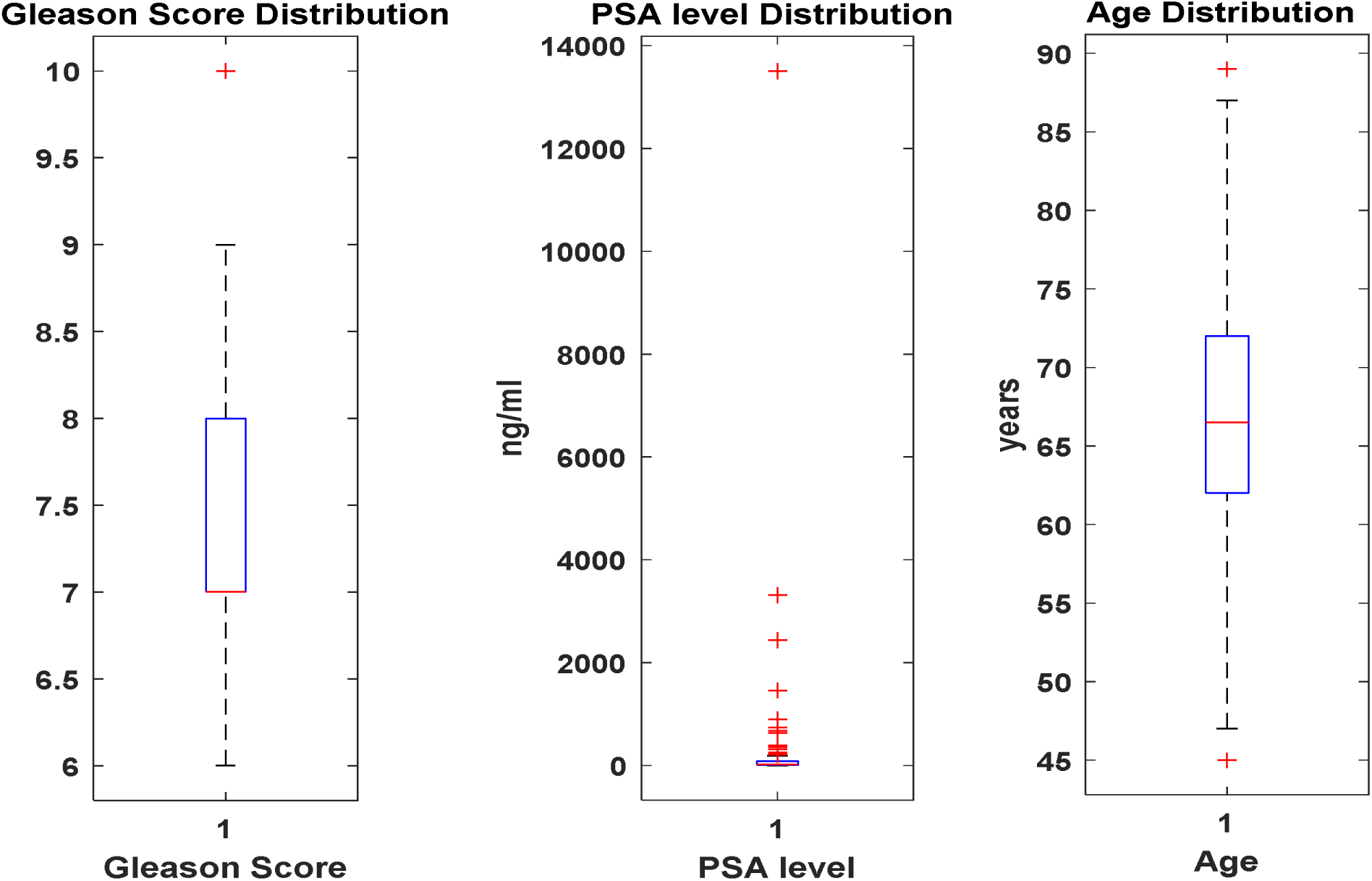
Boxplot representation of Gleason score, PSA level (ng/ml), and age (years) distribution in prostate cancer patients

The frequency and percentage of key attributes found in these Ga-68 PSMA PET/CT reports are given in **Table-2**. Attributes have been categorized based on reporting frequency such as: Highly Reported (≥90%), Moderately Reported (50–89%) and Underreported (<50%) is given in **Table-3**. The RAS for each report is shown both as line plot and boxplot in **Figure-2**. Bar chart of each attribute having its values across 200 reports demonstrating in how many reports each attribute were observed in shown in **Figure-3**. Reporting Adequacy Score varied from a minimum of 31% to a maximum of 78%. Based on the interpretation criteria defined for RAS, 91% (182 out of 200) of the reports have 50-80% RAS that is the report is partially adequate, with notable missing elements; and 9% (18 out of 200) reports have RAS less than 50% indicating the report is inadequate and may not meet clinical needs effectively. These missing attributes include Lesion SUVmax, Neurovascular bundle involvement, Levator ani muscle involvement, Regional lymph node SUVmax, Non-regional lymph node SUVmax, Bone metastasis lesion count, and Bone metastasis SUVmax. In contrast, some attributes, such as the presence or absence of non-regional lymph nodes and the presence or absence of bone metastases, were documented in all reports.

**Figure 2:**
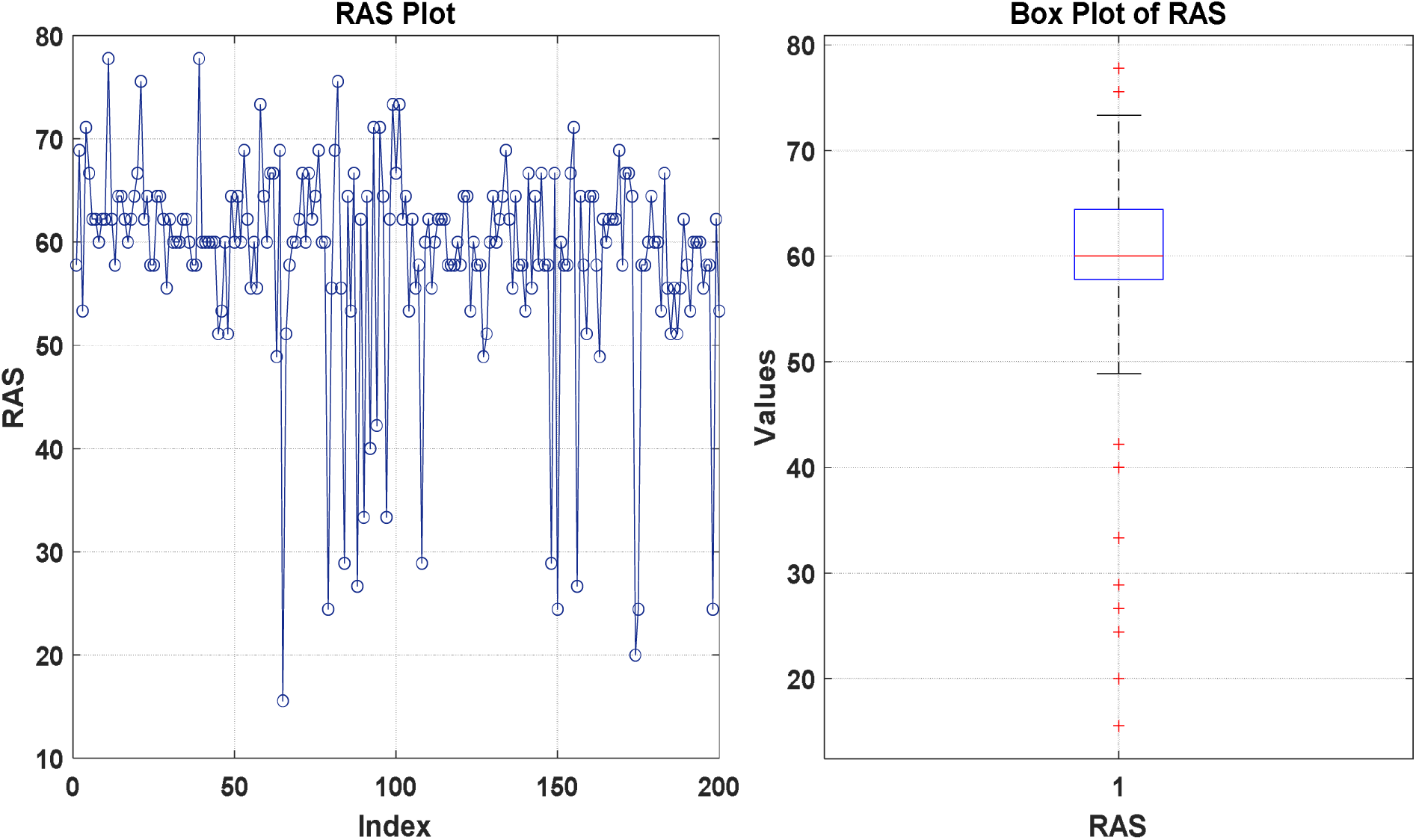
Line plot (left) and boxplot (right) of Report Adequacy Score (RAS). The line plot shows variations in RAS for each report, while the boxplot summarizes the distribution, highlighting the median, interquartile range, and outliers for each report

**Figure 3:**
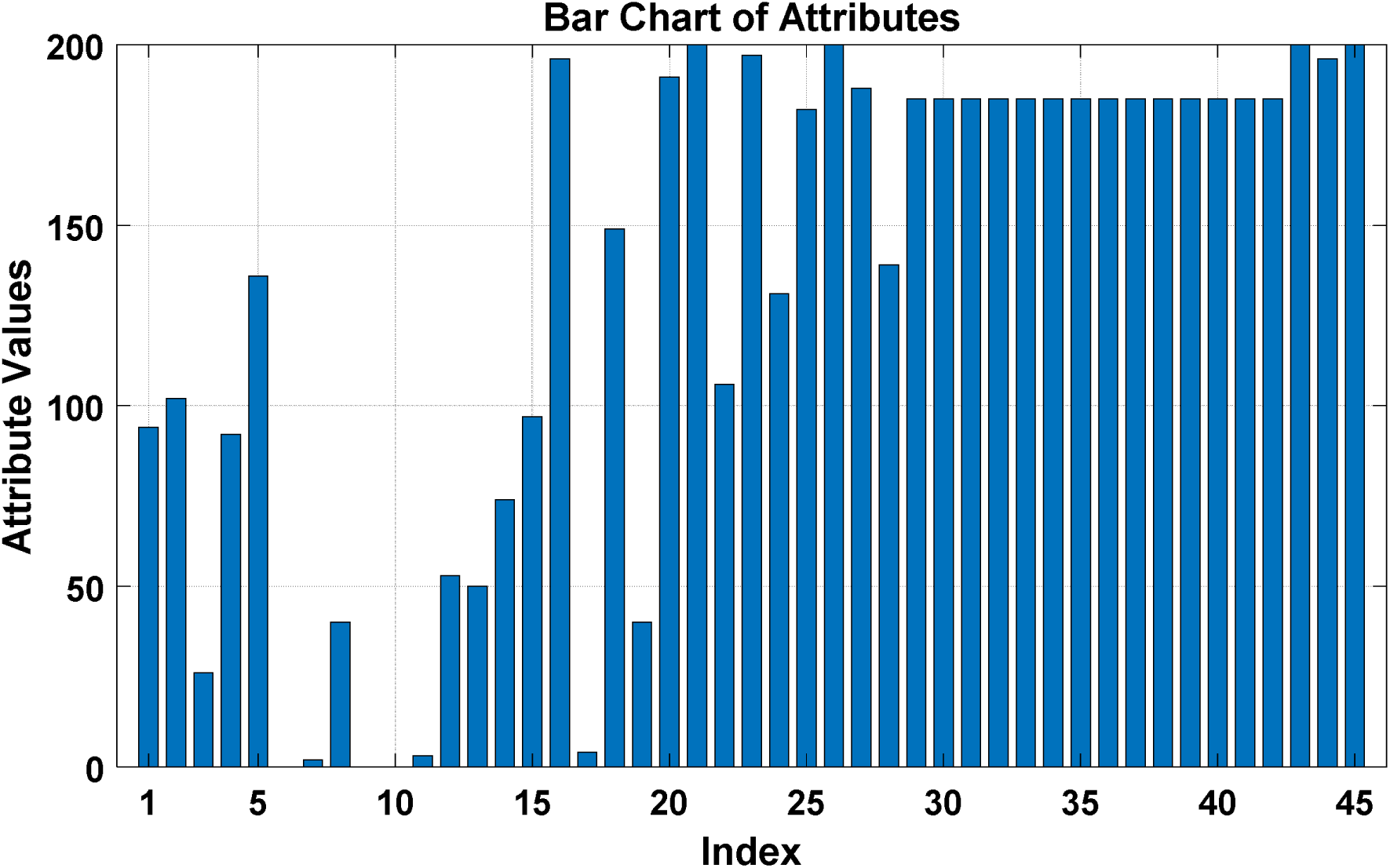
Bar chart showing the values of each attribute across 200 reports. The height of each bar represents the attribute’s value, helping to visualize differences and trends in the dataset. Since this study evaluated reports of patients who underwent Ga-68 PSMA PET/CT imaging solely for baseline evaluation (as per the inclusion criteria), the 46^th^ attribute, ”Comparison (if follow-up scan, compare with previous findings),” is marked as ”N/A.”

### To what extent does variability exist in the terminology used?

The terminology used to describe findings on Ga-68 PSMA PET/CT scans for attributes such as lesion zone, lesion side, uptake patterns in prostate lesions, lymph nodes, and seminal vesicle invasion was inconsistent.

1. The lesion zone was described with specific terminology in 39.5% of the reports, whereas non-specific terms such as ”Entire Gland,” ”Almost Entire Gland,” and ”Entire Prostatic Parenchyma” were used in 7.5% of the reports.
2. When describing the lesion side, specific regions were mentioned in 41% of the reports, while non-specific terms were used in 10%.
3. The absence of tracer uptake in seminal vesicles was described in nine different ways (italicized in **Table-4**), sometimes conveyed separately and sometimes combined with additional details. For instance, the phrase “Seminal vesicles appear bulky with no tracer uptake” merges structural information with uptake findings.

Similarly, inconsistent terminology was observed in other descriptions, such as:

- Terms indicating tracer uptake in seminal vesicles
- Terminology for increased lesion uptake patterns
- Terminology for normal uptake patterns
- Terminology for low lesion uptake patterns

Inconsistencies were also observed in reporting the following attributes: prostate gland size, lesion size, and lymph node size. The prostate gland size was documented in one, two, or three dimensions, and in some instances, it was accompanied by qualifiers such as ”enlarged” or ”prostatomegaly.” Similarly, the size of prostate lesions and lymph nodes was reported either quantitatively, qualitatively, or using both quantitative and qualitative descriptions (**Table-5**).

The variability index of the attributes (in which variability in terminology was observed) and its interpretation is given in **Table- 6**.

### Is the structure and content of the report optimized for seamless AI processing?

A significant number of attributes—22 in total (6 moderately reported and 16 underreported) out of 46 attributes—fall into the moderately to underreported categories (Table 3). An AI model extracting these attribute values would record them as missing. Additionally, the Variability Index (VI) of attributes ranges from 4% to 67% (Table 6), increasing the risk of AI either failing to capture these attributes, leading to missing data, or misinterpreting them, resulting in incorrect information. Consequently, an AI model trained on these 200 reports is likely to exhibit compromised performance.

**Table 2:** Frequency and percentage of key attributes reported in 200 retrospective Gallium-68 PSMA PET/CT scan reports for prostate cancer.

| **Attribute** | **Frequency (out of 200)** | **Percentage (%)** |
| --- | --- | --- |
| Lesion Zone | 94 | 47 |
| Lesion side | 102 | 51 |
| Lesion clock position | 26 | 13 |
| Lesion apex free or not | 92 | 46 |
| Lesion pattern | 136 | 68 |
| Lesion SUVmax | 00 | 00 |
| Prostate Gland size (3D, cm) | 02 | 1 |
| Lesion size (3D, cm) | 40 | 20 |
| Capsular invasion | 00 | 0 |
| Seminal Vesicle invasion (Location, Extent of uptake) | 00 | 00 |
| Neurovascular bundle involvement | 03 | 1.5 |
| Levator ani muscle | 53 | 26.5 |
| Pelvic wall invasion | 50 | 25 |
| Bladder base involvement | 74 | 37 |
| Rectum involvement | 97 | 48.5 |
| Regional Lymph node Presence/Absence | 196 | 98 |
| Regional Lymph node Name | 04 | 2 |
| Regional Lymph node SUVmax | 149 | 74.5 |
| Regional Lymph node Side | 40 | 20 |
| Number of Regional Lymph node | 191 | 95.5 |
| Regional Lymph node Size | 200 | 100 |
| Non-regional Lymph node Presence/Absence | 106 | 53 |
| Non-regional lymph node Name | 197 | 98.5 |
| Non-regional lymph node SUVmax | 131 | 65.5 |
| Non-regional lymph node Side | 182 | 91 |
| Number of non-regional lymph node | 200 | 100 |
| Non-regional lymph node Size | 188 | 94 |
| Bone metastasis Presence/Absence | 139 | 69.5 |
| Bone metastasis Location | 185 | 92.5 |
| Bone metastasis number of lesions | 185 | 92.5 |
| Bone metastasis SUVmax | 185 | 92.5 |
| Visceral Liver metastasis | 185 | 92.5 |
| Visceral Liver metastasis Size in 3D | 185 | 92.5 |
| Visceral Liver metastasis SUVmax | 185 | 92.5 |
| Visceral Liver metastasis, No of lesions | 185 | 92.5 |
| Visceral Lung metastasis | 185 | 92.5 |
| Visceral lung metastasis Size in 3D | 185 | 92.5 |
| Visceral Lung metastasis SUVmax | 185 | 92.5 |
| Visceral Lung metastasis No of lesions | 185 | 92.5 |
| Visceral other organ metastasis | 185 | 92.5 |
| Visceral other organ metastasis Size in 3D | 185 | 92.5 |
| Visceral other organ metastasis SUVmax | 185 | 92.5 |
| Visceral other organ metastasis No of lesions | 200 | 92.5 |
| Urinary tract Physiological uptake | 196 | 100 |
| Urinary tract Physiological uptake unusual distribution | 200 | 98 |

**Table 3:**
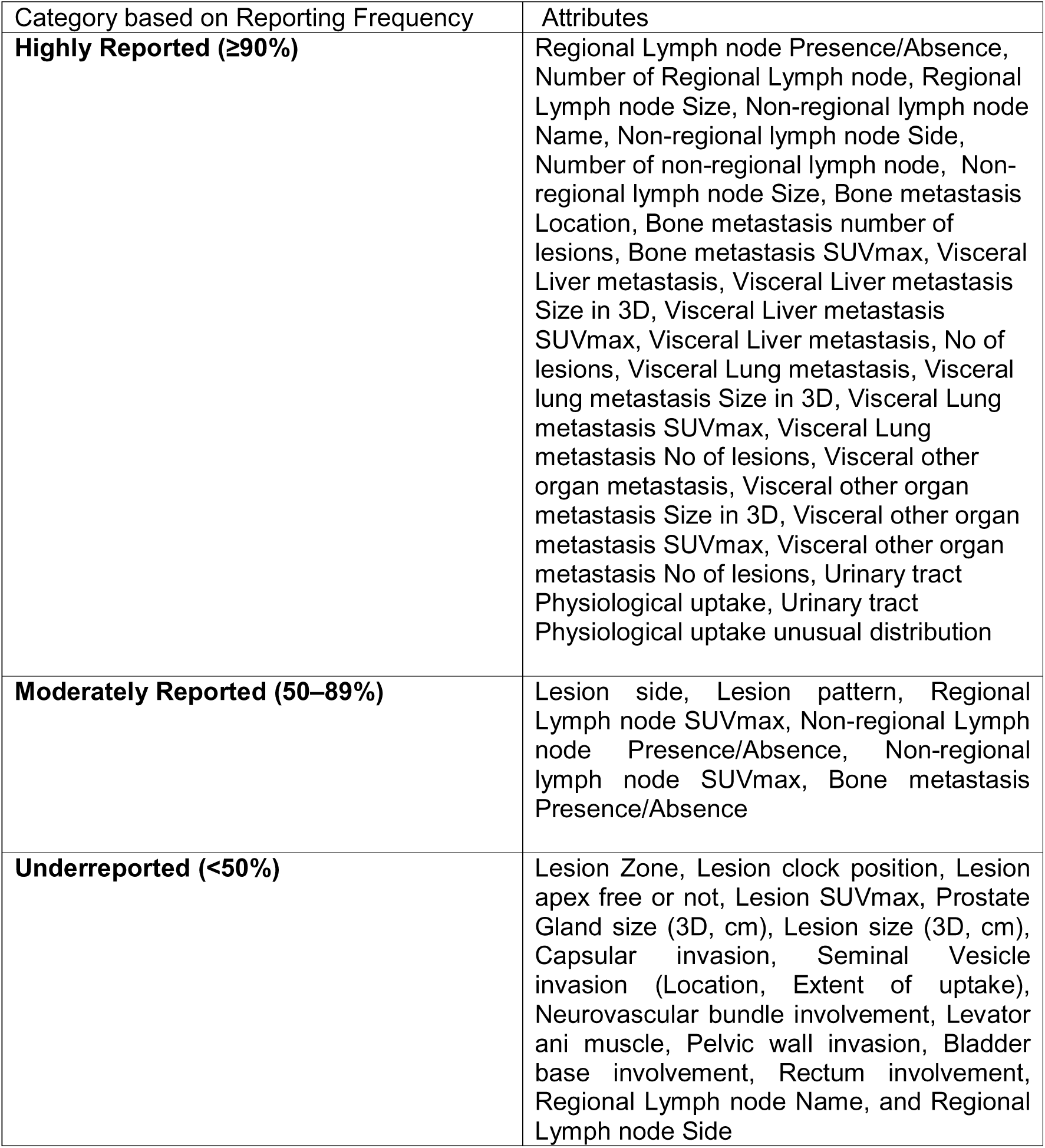
Categorizes the attributes based on their reporting frequency into three groups: Highly Reported (≥90%), Moderately Reported (50–89%), and Underreported (<50%).

**Table 4:**
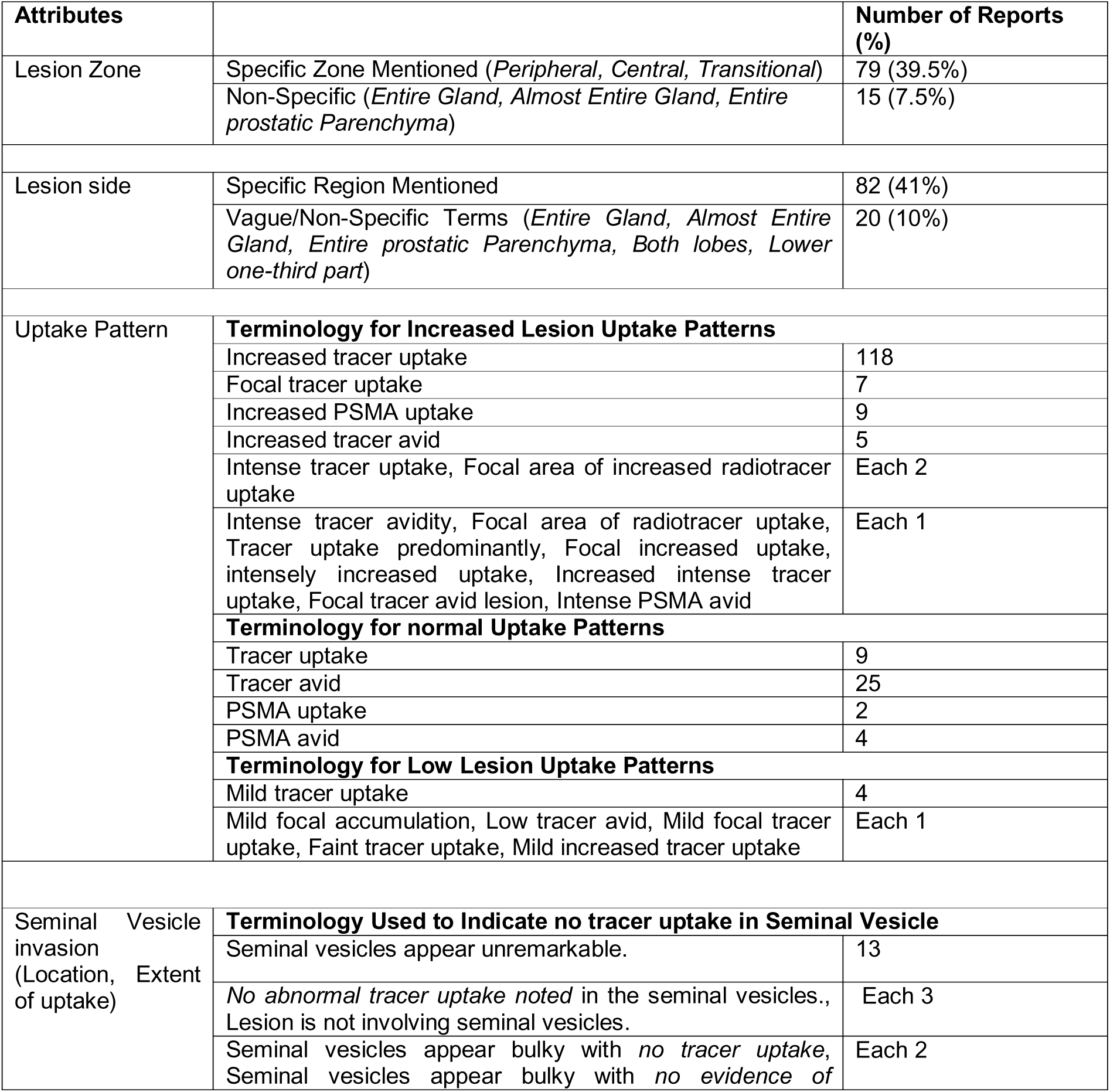
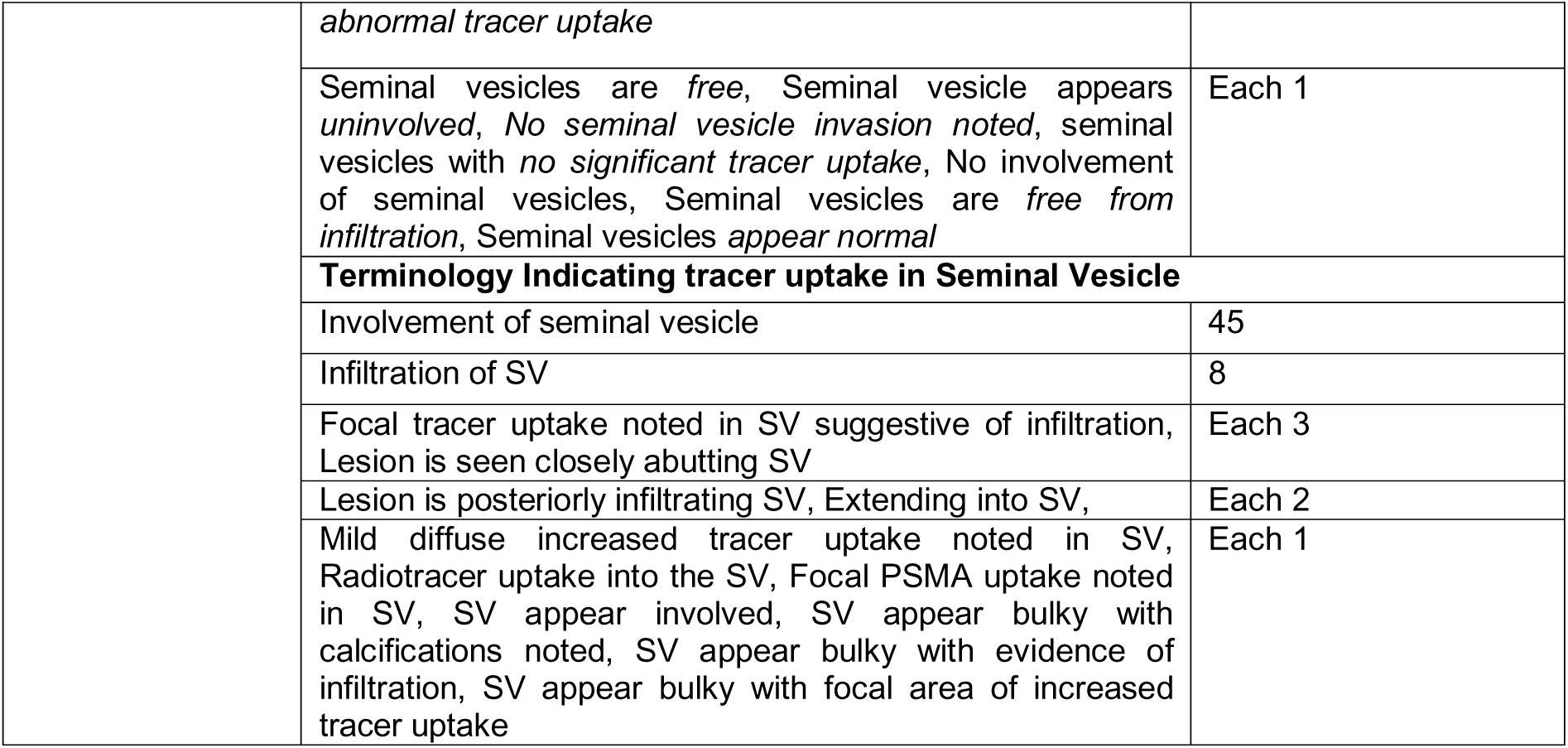
The variability in terminology used to describe lesion characteristics on Ga-68 PSMA PET/CT scans, including lesion zone, lesion side, uptake patterns, and seminal vesicle invasion.

**Table 5:**
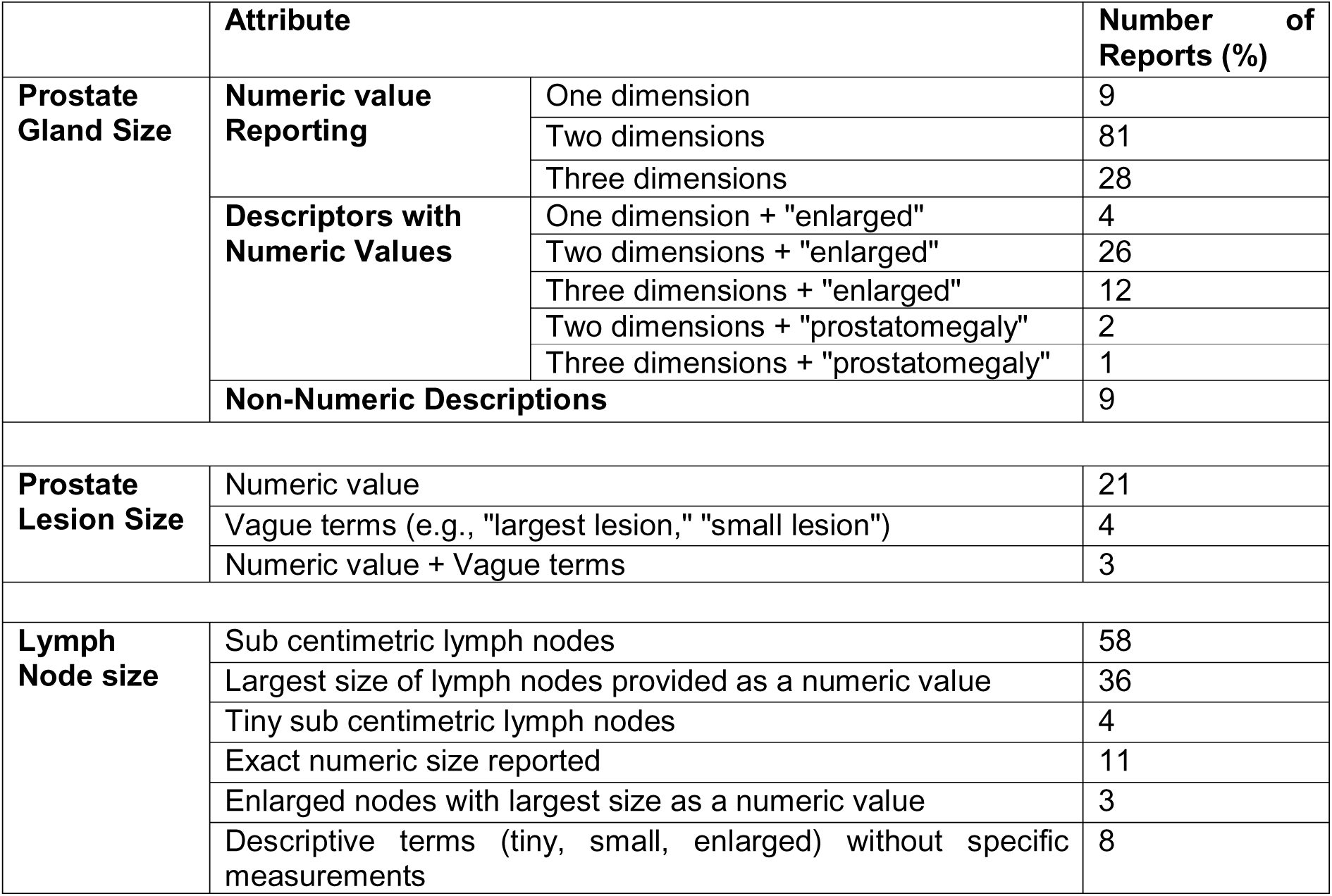
Reporting patterns of prostate gland, prostate lesion, and lymph node sizes in reports, categorizing them into numeric, qualitative, and combined descriptive formats.

**Table 6:** Variability index of different attributes related to prostate and lymph node assessments, categorizing them into minimal, moderate, or high variability based on terminology differences.

| Attribute | Variability Index | Interpretation |
| --- | --- | --- |
| Lesion Zone | 4.25 % | Minimal variability |
| Lesion Side | 5.89 % | Minimal variability |
| Increased uptake pattern | 9.27 % | Minimal variability |
| Normal Uptake pattern | 10 % | Moderate variability |
| Low Uptake pattern | 66.67 % | High variability |
| No tracer uptake in Seminal Vesicle | 40 % | High variability |
| Presence of tracer uptake in Seminal Vesicle | 18.5 % | Moderate variability |
| Lymph Node Size | 5 % | Minimal variability |
| Prostate lesion size | 10.7 % | Moderate variability |
| Prostate gland size | 5.2 % | Minimal variability |

## Discussion

Our study analysed 200 Ga-68 PSMA PET/CT reports (January 2020–June 2024) to assess alignment with clinical needs, terminology variability, and AI suitability. The RAS ranged from 31% to 78%, with 91% of reports scoring between 50–80%, indicating partial adequacy but notable omissions. Key missing attributes included Lesion SUVmax, Neurovascular bundle involvement, Regional and Non-regional lymph node SUVmax, and Bone metastasis details. While some elements, like the presence of non-regional lymph nodes and bone metastases, were consistently reported, terminology inconsistencies were widespread. For instance, lesion zones were precisely described in only 39.5% of reports, while vague terms appeared in 7.5%. Additionally, seminal vesicle involvement was described using nine different phrasings. These findings underscore the need for structured reporting to improve clarity, reduce variability, and enhance both clinical decision-making and AI integration.

The omission of key details in reports is likely due to the time-consuming process of extracting attributes such as prostate gland size (3D), lesion size (3D), SUVmax, and lymph node, bone, and visceral metastasis characteristics from PET images. For instance, measuring SUVmax requires drawing a region of interest, recording values, and manually entering them into the report, while obtaining 3D measurements and counting multiple lesions further adds to the complexity [9]. The reason for missing these quantitative data might also be due to limited numbers of qualified manpower and the number of terminals to process PET images to cater the need of large volume of patients per day our facility is handling. There are also other factors that can be attributed to missing details: software and workflow limitations in PACS and reporting systems may make manual data entry cumbersome; Subjectivity in reporting also plays a role, as certain parameters may not always be deemed essential for immediate clinical decision-making; Variability in imaging protocols or scan quality may lead to cases where some parameters are difficult to assess; The level of training and experience of reporting physicians further influences documentation, with less experienced residents potentially overlooking certain parameters; in routine clinical practice, findings that do not alter patient management may be deliberately omitted to streamline reporting; and the need for efficiency in high-volume nuclear medicine departments may exert pressure on physicians to focus on the most relevant details, leading to selective documentation.

The findings of our study regarding missing information due to the free-text nature of sub- section reporting in semi-structured reports align with the observations made by Goldenberg et al. [10] The partial fulfilment of clinical requirements in Ga-68 PSMA PET/CT reports observed in our study highlights a significant gap in standardized reporting and reinforces the need for structured reporting. Research has demonstrated that standardized imaging frameworks, such as the Prostate Imaging-Reporting and Data System (PI-RADS) for MRI, enhance diagnostic accuracy and interobserver agreement in prostate cancer detection [11]. The absence of a comparable standardized system for Ga-68 PSMA PET/CT imaging may be a key factor contributing to the deficiencies in meeting clinical needs.

Extracting information from the reports was challenging and time-consuming due to the inconsistent and vague terminology used for certain attributes and their values. This lack of uniformity led to ambiguity in clinical interpretation and posed significant challenges in data aggregation. The inconsistency in reporting highlights a major obstacle for AI in extracting key information, ultimately affecting its performance in clinical applications. Standardized structured reporting is essential for enhancing AI-driven analysis, ensuring accurate and consistent interpretations, and improving clinical relevance. Variability in reporting can impact treatment decisions, staging accuracy, and disease monitoring while also limiting AI’s ability to identify meaningful patterns for automated staging and prognostication [12].

Several studies have highlighted the benefits of structured reporting (SR) in oncologic imaging, emphasizing its role in improving report quality, completeness, and communication with clinicians. Leithner et al. surveyed 200 radiologists from 51 countries and found strong support for SR, though adoption in Europe remains below international standards [13]. Marcovici et al. demonstrated that structured chest radiograph reports were completer and more effective than unstructured ones [14]. Other studies have proposed structured templates and compared free-text versus structured reporting in various imaging modalities, including FDG, DOTANOC, and PSMA PET/CT. Niederkohr et al. stressed the need for standardized PET/CT reports, ensuring clarity in clinical history, imaging technique, and findings [15]. Werner et al. introduced structured frameworks like PSMA-RADS, PROMISE, and the EANM Consensus Paper, which enhance prostate cancer imaging accuracy, improve interobserver agreement, and support clinical decision-making. Collectively, these studies highlight the importance of structured reporting in ensuring consistency, reducing errors, and optimizing AI-driven data extraction [16].

Studies consistently highlight the variability in Ga-68 PSMA PET/CT reporting, which hinders both clinical decision-making and AI-driven automation. Structured frameworks like PROMISE and PSMA-RADS offer standardized criteria that improve interobserver agreement, enhance staging accuracy, and facilitate AI training by providing well-defined data categories [17,18]. Yasmin et al. demonstrated the clinical utility of the PSMA score based on PROMISE criteria, though broader validation is needed [19]. Similarly, Esfahani et al. developed a standardized PSMA PET/CT reporting template to improve clarity and clinical communication [20]. Despite these advancements, a universally accepted reporting template remains elusive.

The significance of this study lies in its effort to enhance the clinical utility of Ga-68 PSMA PET/CT by addressing inconsistencies in reporting structure and terminology, which can impact clinical decision-making and AI integration. By systematically analysing reports from a high-volume tertiary care center, this research identifies gaps in the completeness of clinical information and inconsistencies in terminology. The findings highlight the need for structured reporting, which can improve interdisciplinary communication, facilitate AI-driven data extraction, and enhance diagnostic accuracy.

This single-center, retrospective study has limitations, including selection bias from exclusively analyzing baseline evaluation reports, restricting generalizability. While it highlights AI-related challenges, advancements in NLP may address some issues. Additionally, the study does not assess how reporting variability affects clinical outcomes. To overcome these limitations, future work will focus on converting semi-structured Ga-68 PSMA PET/CT reports into a structured format using NLP- and rule-based techniques. Expanding the database to include all report types will enhance AI integration, improve consistency, and support structured reporting in clinical practice.

## Conclusion

The majority of Ga-68 PSMA PET/CT reports were partially adequate, with significant missing details and considerable variability in terminology. The variability was evenly distributed across minimal, moderate, and high levels. Training an AI model on such reports would likely result in slower learning and compromised performance.

## Data Availability

All data produced in the present work are contained in the manuscript

